# Differentiating Benign and Hepatocellular Carcinoma Cirrhotic Nodules: Radiomics Analysis of Water Restriction Patterns with Diffusion MRI

**DOI:** 10.1101/2024.12.08.24318637

**Authors:** Arvin Arian, Fardin Samadi Khoshe Mehr, Babak Setayeshpour, Sina Delazar, Azin Nahvijou, Mohsen Nasiri-Toosi, Maryam Fotouhi

## Abstract

**Objectives:** Current study aimed to investigate radiomics features derived from two-center diffusion-MRI to differentiate benign and hepatocellular carcinoma (HCC) liver nodules.

**Methods:** A total of 328 patients with 517 LI-RADS 2-5 nodules were included. MR images were retrospectively collected from 3T and 1.5T MRI vendors. Lesions were categorized into 242 benign and 275 HCC based on follow-up imaging for LR-2,3 and pathology results for LR4,5 nodules, and randomly divided into training (80%) and test (20%) sets. Preprocessing included resampling and normalization. Radiomics features were extracted from lesion volume-of-interest (VOI) on diffusion Images. Scanner variability was corrected using ComBat harmonization method followed by High-correlation filter, PCA filter, and LASSO to select important features. Best classifier model was selected by 10-fold cross-validation, and accuracy was assessed on the test dataset.

**Results:** 1,434 features were extracted, and subsequent classifiers were constructed based on the 16 most important selected features. Notably, support-vector machine (SVM) demonstrated better performance in the test dataset in distinguishing between benign and HCC nodules, achieving an accuracy of 0.92, sensitivity of 0.94, and specificity of 0.86.

**Conclusions:** Utilizing diffusion-MRI radiomics, our study highlights the performance of SVM, trained on lesions’ diffusivity characteristics, in distinguishing benign and HCC nodules, ensuring clinical potential. It is suggested that further evaluations be conducted on multi-center datasets to address harmonization challenges.

**Advances in knowledge:** Integration of diffusion radiomics, for monitoring water restriction patterns as tumor histopathological index, with machine learning models demonstrates potential for achieving a reliable noninvasive method to improve the current diagnosis criteria.

## Introduction

Hepatocellular carcinoma (HCC), 75% of primary liver cancers, is the sixth most frequent cancer worldwide and the fourth leading cause of cancer-related mortalities ^1^. The American College of Radiology has approved the Liver Imaging Reporting and Data System (LI-RADS) to standardize information and report hepatocellular masses in patients with cirrhosis. Based on the tumors’ typical features assessed on CT or MR modalities, this classification reports the relative risk of HCC development in categories ranging from LR-1 (definitely benign) to LR-5 (definitely HCC). Accurate differentiation between benign masses and (HCC), particularly within LR-3 and LR-4 categories, remains challenging, necessitating consideration of histopathological results alongside imaging methods despite the invasive nature of biopsy ^2–6^. Therefore, finding imaging markers associated with a higher risk of progression from intermediate likelihood of HCC to HCC is important.

MRI demonstrates superior sensitivity and specificity compared to CT and ultrasound imaging for early assessment of HCC nodules, enabling early diagnosis in cirrhotic patients’ treatment pathways ^7^. Diffusion-weighted imaging (DWI) is a type of non-invasive functional MRI that assesses water restriction patterns and provides insights into tumor microstructural features like cellular density, cellular proliferation, vascularity, and heterogeneity ^2,3,6^. Conflicting results have been found for the visual features of diffusion MRI in differentiating between LR-3, 4, and 5 ^5,8–17^. Signal hyperintensity on high b-value DWI is associated with a high risk of progression toward HCC^18^, which makes DWI an ideal candidate over other MRI modalities for further investigation. Beside conventional features assessed from DWI, Radiomics, as a promising quantitative method, evaluates morphological features such as shape and texture, offering insights into pathophysiological changes encompassing inter- and intra-tumor heterogeneity at the millimeter scale^6,19–23^.

The performance of radiomics in enriching machine learning algorithms in liver imaging was evaluated in different MRI sequences, including T1, T2-weighted, DWI, and dynamic contrast-enhanced (DCE) perfusion ^13,24–28^. DCE-MRI is a powerful imaging technique for quantifying tumor physiological properties, such as perfusion, vascularity, and permeability, which correlate with tumor aggressiveness. Recent studies demonstrate the DCE-MRI’s potential, particularly when combined with radiomics, in predicting tumor biological features, especially in early detection of HCC nodules ^29–31^. However, multicenter implementation is challenging due to variability in contrast agents, protocol standardization, artifact sensitivity, and quantification complexity. In contrast, DWI offers reproducibility, Non-Invasiveness, and easier standardization, providing reliable method in multicenter studies^32,33^. Many researchers have found that ML models trained on radiomics features extracted from DWI can achieve high accuracy in the early diagnosis and prognosis of HCC. However, these studies still suffer from reliability and validity on external dataset ^13,24^ .

In the current landscape, limited research has explored applying radiomics features of water diffusivity patterns for reliable HCC and benign nodules differentiation. Consequently, a pressing need exists to train a machine learning algorithm using specific features and validate its efficacy on external datasets, ultimately constructing the most precise and reliable diagnosis-aided algorithm for clinical translation. This study addresses this gap by training classifier algorithms utilizing DWI-driven radiomics features and evaluation on external dataset to classify HCC and benign liver nodules effectively.

## Methods

### Patients

This retrospective study was approved by the institutional review board of the Cancer Institute. All patients with cirrhotic nodules who underwent MRI between January 2018 and June 2023, including 1133 patients, were initially included in the study. All cirrhotic individuals over the age of 18 years with at least one liver MR scan were included. Exclusion criteria are depicted in Figure 1 as follows: 1. Patients with no LI-RADS 2–5 liver lesions, 2. patients without a diagnosis through imaging and follow-up (LR-2,3) or biopsy (LR-4, 5), 3. prior transarterial chemoembolization treatment due to variable signal increase in DWI values or interventional surgery before the initial MRI scan, 4) prior malignancy, and 5) low-quality diffusion imaging. Finally, A total of 328 patients with 517 nodules met the inclusion criteria (Table 1).

**Figure 1.**
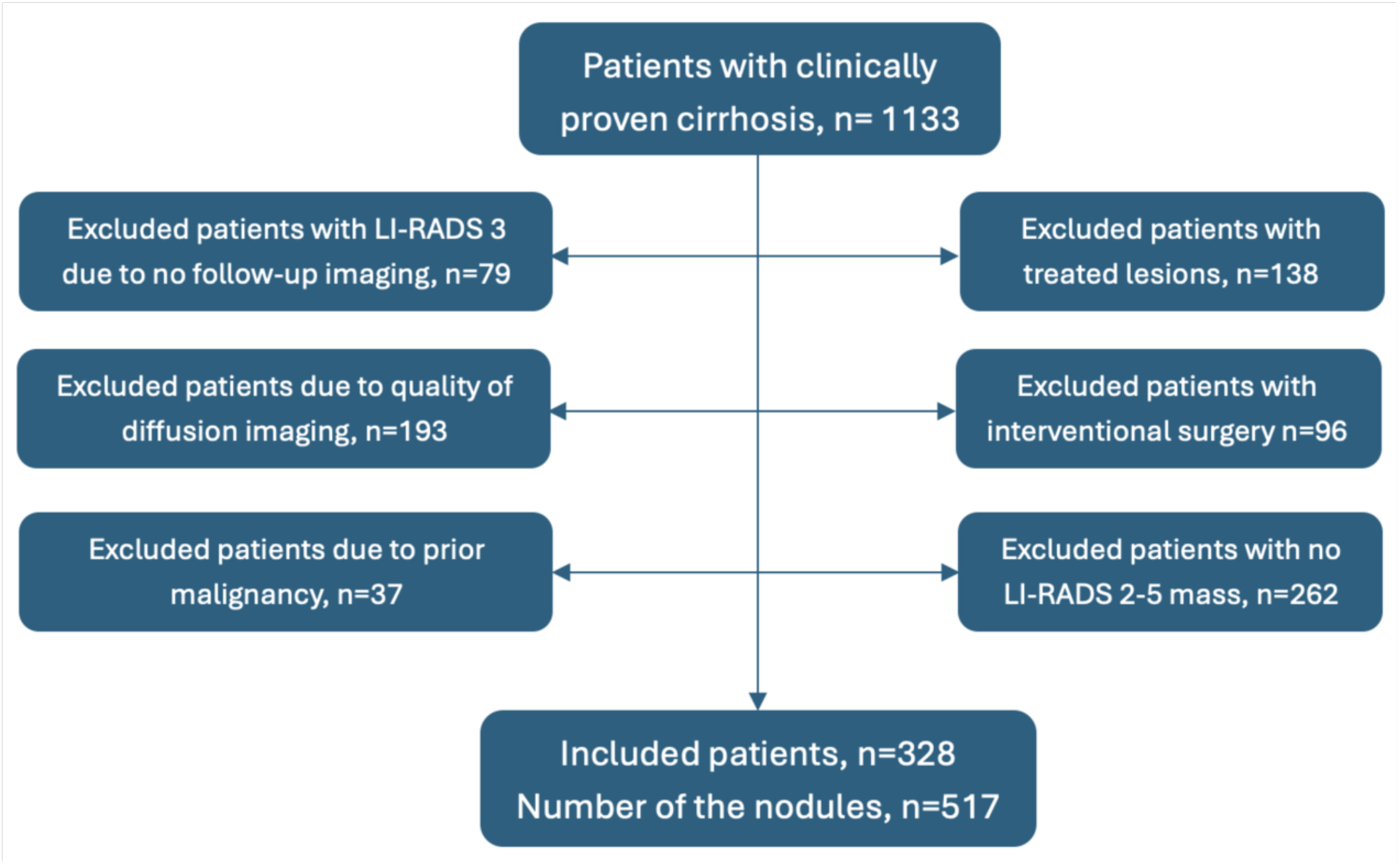
Patient inclusion and exclusion flowchart.

**Table 1.** Study characteristics.

| Parameters | Benign<br>(n=242) | HCC<br>(n=275) | P-value |
| --- | --- | --- | --- |
| LI-RADS 2 | 118 | 7 | - |
| LI-RADS 3 | 75 | 40 | - |
| LI-RADS 4 | 42 | 95 | - |
| LI-RADS 5 | 7 | 133 | - |
| Age | 64.12±9.7 | 69.2±13.1 | 0.62 |
| Sex(M/F) | 181/61 | 173/102 | 0.07 |
| Tumor size (mm) | 19.27±13.99 | 34.65±33.74 | < **0.01** * |
| APHE(Yes/No) | 87/155 | 193/82 | 0.04 |
| Capsule(Yes/No) | 104/138 | 164/111 | 0.72 |
| Washout(Yes/No) | 93/149 | 231/44 | < **0.01** * |
| Lesion ADC (m ± sd) mm<sup>2</sup>/s | 1520±127 | 1384±184 | 0.05 |
| Liver ADC (m ± sd) mm<sup>2</sup>/s | 1207±158 | 1213±161 | 0.46 |
| Paravertebral ADC (m ± sd) mm<sup>2</sup>/s | 1585±189 | 1591±133 | 0.29 |
| ADC1 ratio (lesion/liver) | 1.26±0.28 | 0.92±0.21 | 0.05 |
| ADC2 ratio (lesion/paravertebral) | 0.92±0.17 | 0.85±0.12 | 0.02 |

Arterial phase hyperenhancement: APHE, Hepatocellular Carcinoma: HCC, ADC: Apparent Diffusion Coefficient, LI-RADS: Liver Imaging Reporting and Data System. \* p < 0.01

All patients with LR-4,5 lesions or those considered for a liver transplant underwent nodule biopsy (277 pathology-proven nodules). Follow-up imaging was performed on LR-2,3 nodules for a minimum of 12 months. We divided all nodules into two cohorts: 1- benign nodules (242 nodules; 118 nodules from LR-2, 75 nodules from LR-3, 42 nodules from LR-4, and 7 nodules from LR-5) 2- HCC nodules (275 nodules; 133 nodules from LR-5, 95 nodules from LR-4, 40 nodules from LR-3, and 7 nodules from LR-2).

### Image Acquisition

MRI was performed using 3T (GE Medical System, Discovery 750w) and 1.5T (GE Medical System, Signa) MRI machines. The routine modalities and protocols included the following: 1) DWI with three orthogonal directions along with respiratory-triggered 2D echo planar imaging (EPI) with three b-values (0, 200, 800 s/mm^2^) was used before Gadolinium contrast agent injection. ADC maps were calculated in the prepared workstation. 2) DCE-MRI, LAVA sequence, which is based on a 3D spoiled gradient-recalled echo (SPGR) sequence with uniform fat suppression, was performed in the axial plane with a breath-hold 2D gradient echo T1-weighted sequence. Data acquisition began 10 seconds before injecting the contrast agent to acquire the baseline signal intensity. Thirty-six dynamic contrast slices with a temporal resolution of 4 s/image were acquired during 66 acquisition phases and normal breathing 3) Axial T2-weighted with single-shot fast spin echo (SSFSE). Detailed information about the imaging protocol is listed in Table 2.

**Table 2.**
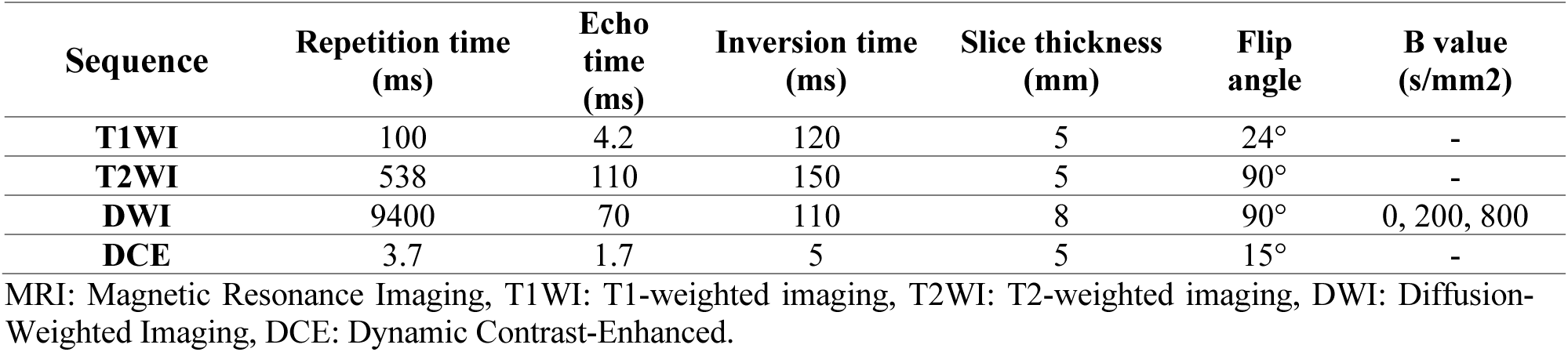
MR Imaging parameters.

| Sequence | Repetition time<br>(ms) | Echo<br>time<br>(ms) | Inversion time<br>(ms) | Slice thickness<br>(mm) | Flip<br>angle | B value<br>(s/mm<sup>2</sup>) |
| --- | --- | --- | --- | --- | --- | --- |
| T1WI | 100 | 4.2 | 120 | 5 | 24° | - |
| T2WI | 538 | 110 | 150 | 5 | 90° | - |
| DWI | 9400 | 70 | 110 | 8 | 90° | 0, 200, 800 |
| DCE | 3.7 | 1.7 | 5 | 5 | 15° | - |

MRI: Magnetic Resonance Imaging, T1WI: T1-weighted imaging, T2WI: T2-weighted imaging, DWI: Diffusion-Weighted Imaging, DCE: Dynamic Contrast-Enhanced.

### Data labeling, Image preprocessing, and lesion segmentation

Two radiologists with 8 and 12 years of diagnostic MR imaging experience assessed the patients’ MR images and established a consensus using LI-RADS v2018 for primary LI-RADS 2–5 categorization ^34^. The kappa score for inter-rater variability between radiologists was 0.76. In cases of disagreement, the subjects were referred to a supervising radiologist for final decision.

To guarantee both reproducibility and reliability, we followed the image biomarker standardization guidelines (ref1). MRI images were resampled to a uniform voxel size of 2x2x2 mm³. Z-score normalization was then applied, transforming pixel intensities to have a mean of 0 and a standard deviation of 1. Linear scaling was then used to map the z-scores to a display range of [0, 255] for clear visualization.

Manual segmentation on lesions was performed using ITK-SNAP software. The boundary of lesions was drawn on axial DWI images (Figure 2). Segmentation was performed by the first radiologist and checked by the senior radiologist to ensure the reproducibility and consistency of the results. Felisa Kappa among radiologists was 0.85 in lesion segmentation.

**Figure 2.**
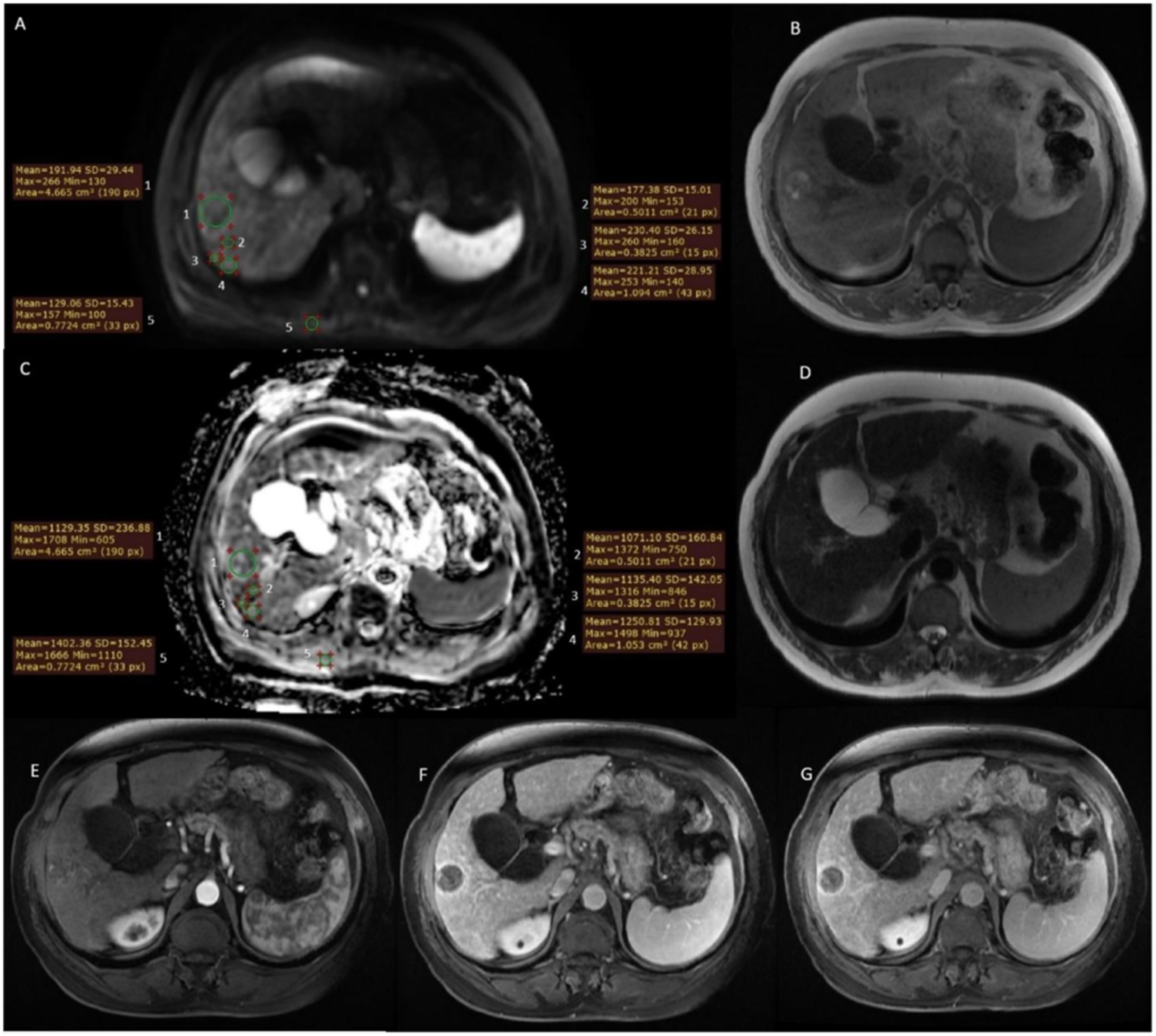
A case with a solitary cirrhotic nodule with hepatocellular carcinoma proven pathology. Region of interests (ROIs) are placed on ADC map for the nodule, liver, and paravertebral muscle (A and C). ROIs are positioned for lesion ADC value on each slice. Liver ADC is calculated with three ROIs placed at the surrounding liver avoiding bile ducts and vessels. The mean value for liver ADC was added. ROI for Paravertebral ADC was placed on the right paravertebral muscle. On DWI (b value, 800 s/mm^2^), restriction is seen (A) along with an ADC map with low signal areas corresponding to restriction in the tumor (C). The T1-weighted image reveals a heterogeneous hyperintensity in the lesion due to fat-containing (B) with mild hyperintensity in the T2-weighted slice (D). The nodule demonstrates arterial phase hyperenhancement (E) with washout in the portal venous phase (F) and enhancing capsule in the delayed phase (G).

### Feature Extraction and selection

Following 3D segmentation of the lesions, radiomics features were extracted from DWI images using PyRadiomics 3.0.1, an open-source Python library, which computes seven distinct feature categories: first-order statistics, shape-based, grey level co-occurrence matrix (GLCM), gray level size zone matrix (GLSZM), gray level run length matrix (GLRLM), gray level dependence matrix (GLDM), and neighboring gray-tone difference matrix. The 16 form descriptors and features extracted from the original and derived images were adjusted for PyRadiomics (LoG with five sigma levels, one level of wavelet decomposition yielding eight derived images, and images derived using square, square root, logarithm, and exponential filters) ^35^.

To adjust for scanner-related variability in radiomics features from 1.5T and 3T DWI images, ComBat was applied using the parametric model. This approach corrects for batch effects by directly modeling and adjusting for scanner-induced variability based on predefined parametric assumptions. The implementation, which differs from the standard EB approach, was chosen to meet specific study requirements. R scripts for ComBat are available at https://github.com/Jfortin1/neuroCombat_Rpackage/.

The number of retrieved characteristics exceeded the number of patients, so feature selection was utilized to avoid overfitting. After extracting 1,434 features, we explored three methods: a high correlation filter, a linear combinations filter, and the 10-fold cross-validation LASSO technique. To remove redundant features, the high correlation filter eliminates variables with substantial absolute correlation, with thresholds of 0.8 adjusted to maintain consistent features. The linear combinations filter, utilizing Principle Coefficient Analysis (PCA) with a 95% variance threshold, addressed collinearity by iteratively removing columns until achieving full rank. These methods yielded 51 features post-filtering. Employing the LASSO technique with 10-fold cross-validation further refined feature dimensionality, ensuring robustness against overfitting. Parameter optimization, including regularization strength, was conducted via cross-validation to optimize model performance. These feature reduction strategies, executed using the scikit-learn 1.5.0 and pandas 2.2.2 libraries in Python.

For the robustness and generalizability of our models, we split our dataset into a training set (414 lesions, 371 lesions from 3T and 43 lesions from 1.5T) for hyperparameters tuning and model selection using 10-fold cross-validation, and an external test dataset (103 lesions, 91 lesions from 3T and 12 lesions from 1.5T) for evaluating the performance of the selected model on unseen data.

### Statistical Analysis

For Model construction, we selected classifiers, including Logistic Regression (LR), Support Vector Machines (SVM), RF, Multilayer perceptron (MLP), and Gradient Boosting (GB), for a comprehensive analysis. Each classifier offers unique advantages and characteristics, making them appropriate candidates for further study. To ensure a robust assessment, we utilized the scikit-learn library in Python, a widely adopted toolkit for machine learning tasks ^36^. The classifiers were trained on the original training set, and their performance was evaluated on both the original and test sets. Evaluation metrics, including accuracy and confusion matrices, were calculated to provide a deep understanding of each classifier’s strengths and weaknesses.

Furthermore, Receiver Operating Characteristic (ROC) curves were employed to assess the classifiers’ ability to differentiate between the two classes across different threshold values. The Area Under the Curve (AUC) was calculated for each ROC curve, providing a quantitative measure of the classifiers’ overall classification performance. The student’s t-test was utilized to compare continuous variables between benign and malignant groups, while the chi-square test was applied to analyze differences in categorical variables across the two groups.

## Results

### Clinical Characteristics

The study included 328 individuals (208 males, mean age 57.08 ± 11.33, range 29–81 years) with 517 lesions (mean size 33.09 ± 30.93 mm) of LI-RADS 2–5. The clinical characteristics of the nodules are shown in Table 2. Arterial phase hyperenhancement was seen in 68% of HCC and only 35% of benign masses; 44% of benign and 58% of HCC lesions contained enhancing capsule, and 82% in HCC and 36% in benign lesions reported a non-peripheral washout appearance. Tumor size and washout from DCE showed significant p-value in differentiation of HCC and benign nodules. The mean ADCs for lesions, liver, paravertebral muscle, ADC1 ratio (lesion to liver) and ADC2 ratio (lesion to paravertebral) for benign and HCC lesions are shown in Table 2.

### Feature selection

The workflow for image processing and analysis is depicted in Figure 3 and includes image preprocessing, lesion segmentation, feature extraction and selection, model creation, and validation. Initially, 1,434 features were extracted from seven radiomics categories. Applying a correlation threshold reduced the number to 150 features. Subsequent PCA analysis, aimed at eliminating collinear features, further reduced this number to 51. Lasso regression identified 17 features with non-zero coefficients. Finally, by applying an optimal lambda threshold of 0.69, 16 features were selected for training the classification models. The results of each step are depicted in Figure 4 and the selected features are listed in Table 3. GLCM, shape, and first-order feature classes showed the highest importance in benign and HCC classification. The top features identified for the GLCM, GLDM, GLSZM, and first-order classes belonged to the Wavelet filter. In the Shape class, the chosen features were Major axis and elongation from original image.

**Figure 3.**
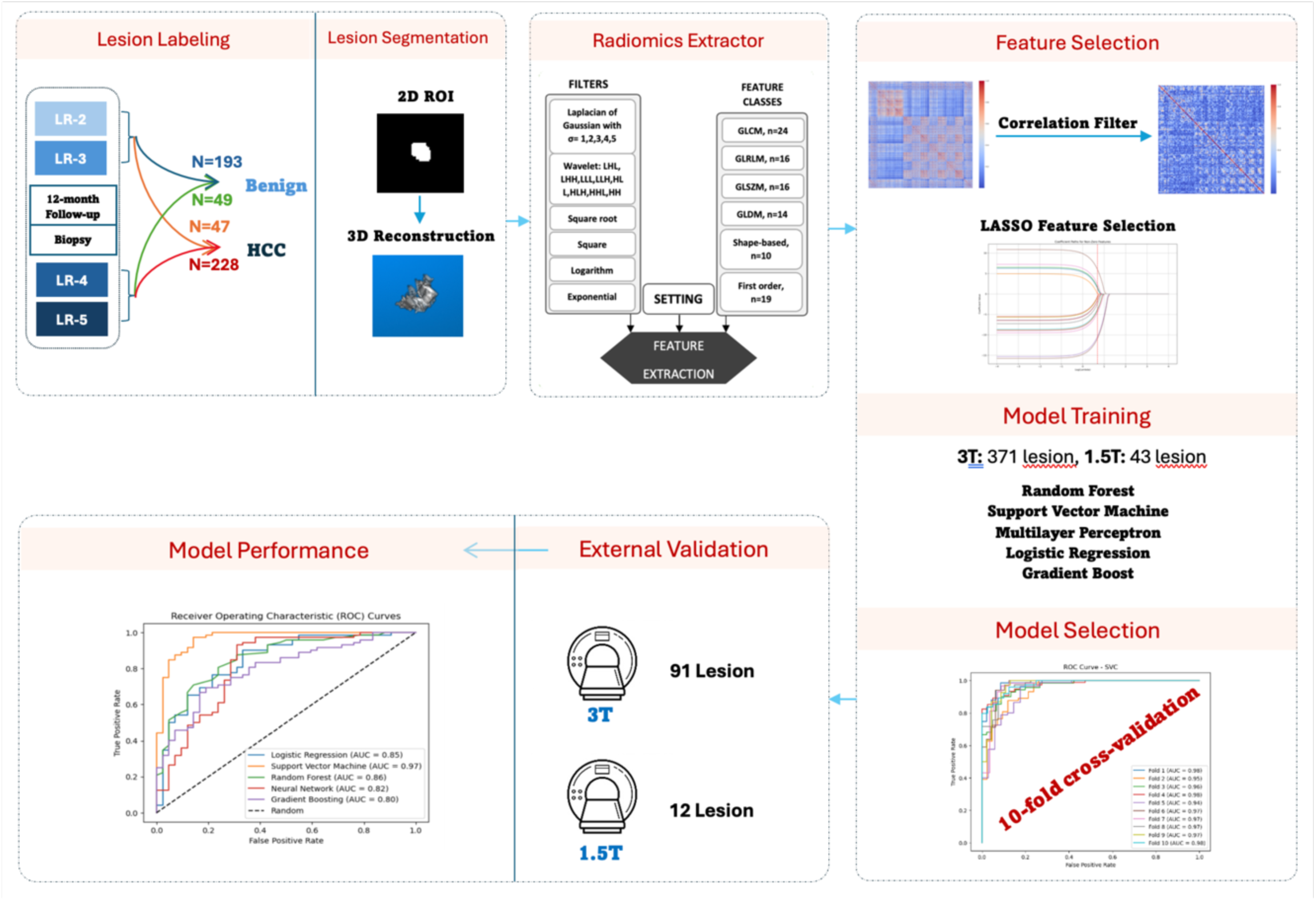
Study workflow and lesion segmentation. DWI and lesion segmentation of cirrhotic nodules (left), Radiomics feature extraction (middle), feature selection, Machine-learning model construction (right), and evaluation with external dataset.

**Figure 4.**
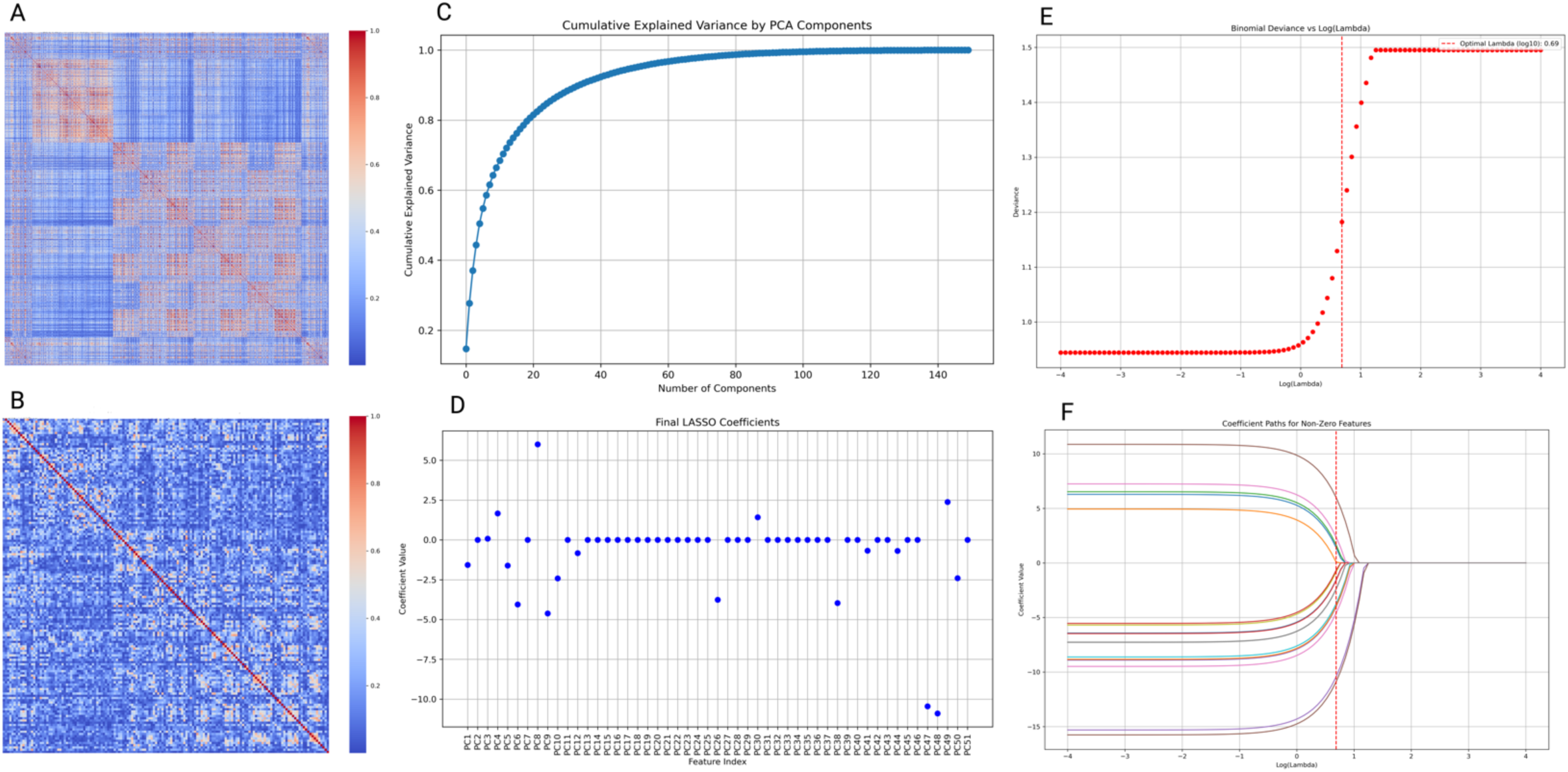
Feature Selection Workflow: (A) Correlation map of all 1,434 radiomic features. (B) Correlation map after applying a high-correlation filter with a 0.8 threshold, reducing the number of features to 150. (C) Principal Component Analysis (PCA) with a 0.95 variance threshold, resulting in 51 features. (D) LASSO regression identified 17 features with non-zero coefficients. (E) The deviance-log(lambda) curve determined a regularization parameter threshold of 0.69. (F) Applying this threshold led to the selection of 16 features.

**Table 3.**
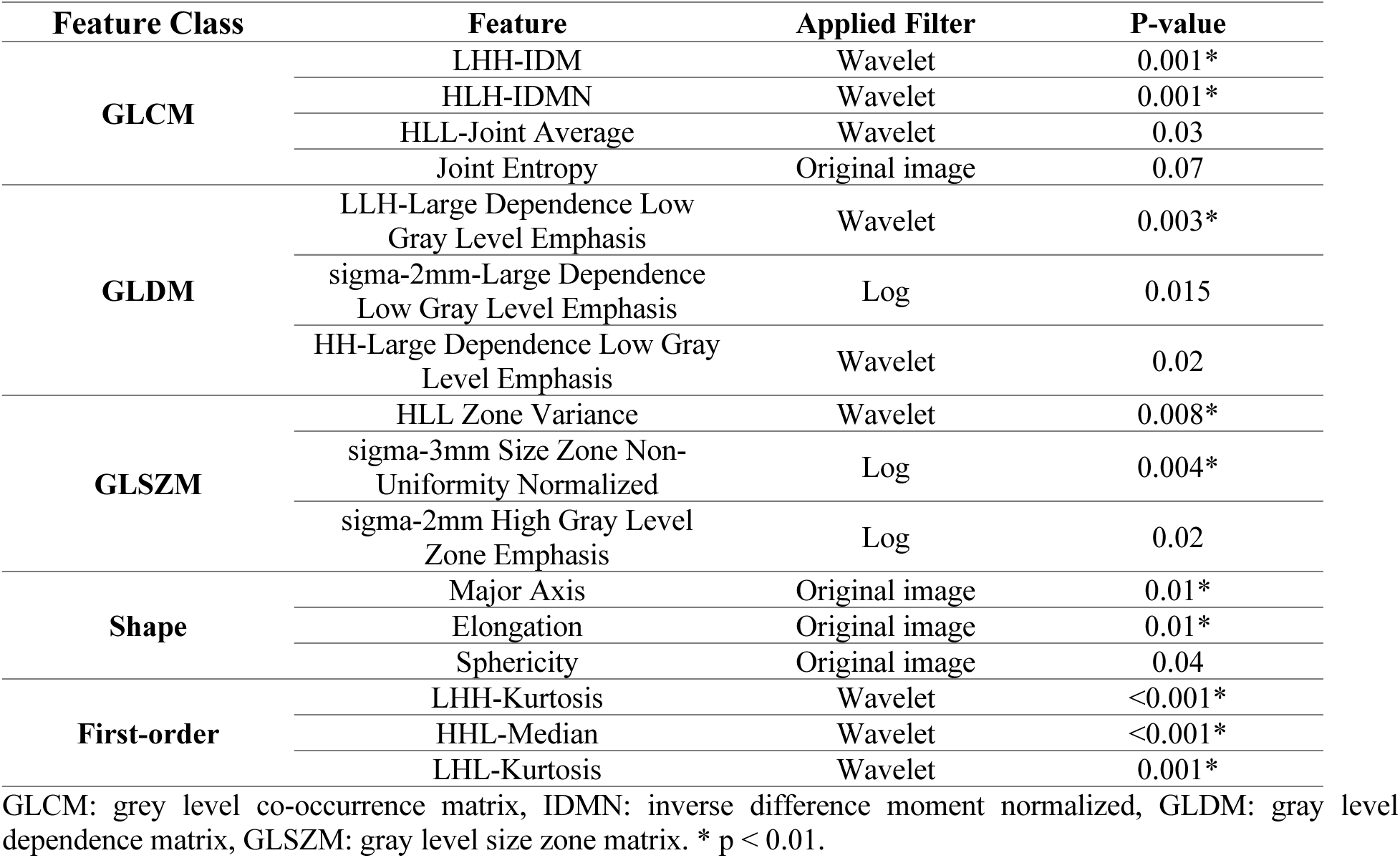
Selected radiomics-driven features in differentiation HCC and benign nodules.

### Model evaluation

Precision and accuracy were employed to assess the radiomics model’s performance, as detailed in Table 4. ROC curves in Figure 5 illustrate the classifier function, with Logistic Regression achieving an accuracy of 0.69, sensitivity of 0.59, specificity of 0.86, and ROC AUC of 0.85. SVM exhibited superior performance with an accuracy of 0.92, sensitivity of 0.94, specificity of 0.86, and ROC AUC of 0.97. RF displayed an accuracy of 0.56, sensitivity of 0.29, specificity of 1.00, and ROC AUC of 0.86. Neural Network yielded an accuracy of 0.45, sensitivity of 0.14, specificity of 0.97, and ROC AUC of 0.82. Gradient Boosting achieved an accuracy of 0.61, sensitivity of 0.40, specificity of 0.90, and ROC AUC of 0.80.

**Figure 5.**
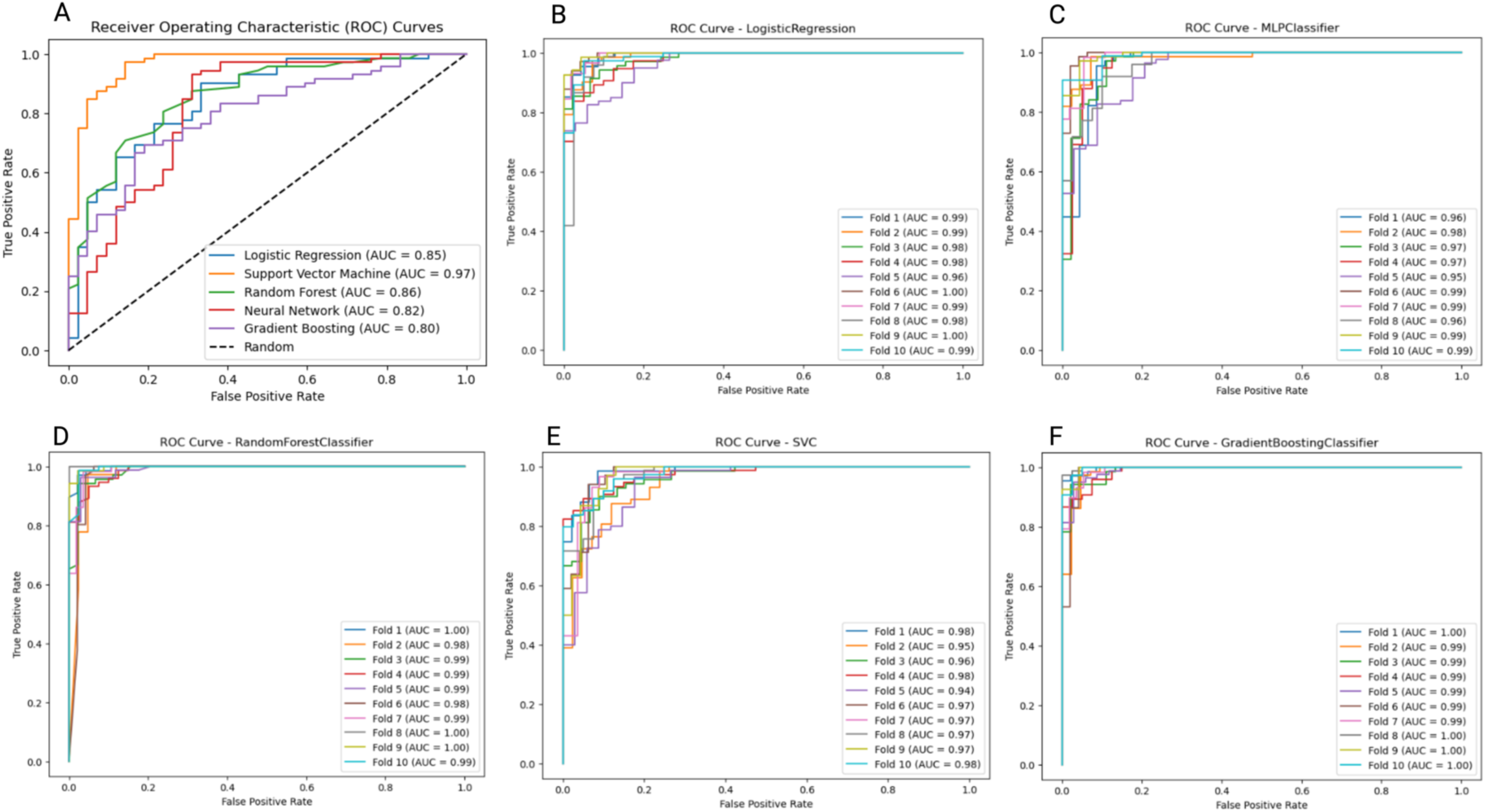
Receiver operating characteristic (ROC) curves. The horizontal axis is the false positive rate, and the vertical axis shows the true positive rate. Different Models’ performance on the external test dataset (A), and training dataset for Logistic Regression (B), Multilayer Perceptron (C), Random Forest (D), Support Vector Machine (E), and Gradient Boosting (F).

**Table 4.** ML models efficiency on external dataset.

| Classification model | Accuracy | Sensitivity | Specificity |
| --- | --- | --- | --- |
| **Logistic Regression** | 0.69 | 0.59 | 0.86 |
| **Support Vector Machine** | 0.92 | 0.94 | 0.86 |
| **Random Forest** | 0.56 | 0.29 | 1.00 |
| **Neural Network** | 0.45 | 0.14 | 0.97 |
| **Gradient Boosting** | 0.61 | 0.40 | 0.90 |

Despite the consistent and high accuracy observed across all models during training, as evident from the ROC curves across various folds shown in Figure 5, a noteworthy divergence becomes apparent when assessing their performance on the test dataset. Only the SVM model maintains its accuracy on the test data, showcasing a robust generalization capability. In contrast, the remaining models experience a decline in accuracy when evaluated on the test dataset.

## Discussion

The differentiation between benign and HCC lesions requires accurate biomarkers for appropriate treatment planning. Due to the similarity in MRI-derived markers between LI-RADS 3, 4, and 5, particularly when comparing 3 with 4 and 5, there is a practical need to investigate novel imaging techniques, characteristics, and classification models ^37,38^. Our initial findings suggest that the water restriction pattern, analyzed through reliable radiomics tools, holds promise in reliably differentiating between benign nodules and HCC.

The basis of current imaging diagnosis of primary cirrhotic nodules, especially HCC, is the use of visual features of contrast-enhanced MRI as a complicated method and in some cases with controversy in the indication ^39–41^. Peng et al. showed that ADC values significantly classify benign and HCC lesions in Asian populations ^11^. Some studies have explored alternative approaches, employing AI and quantitative features for improving LI-RADs categorization accuracy or predicting pathology results. Hai-Feng Liu trained ML models with DCE-MRI radiomics features to classify HCC into well, moderate, and poorly differentiated HCC classes according to pathology results and reached an AUC of 0.86 on the validation dataset ^42^. Rong Hu used radiomics features of T1-contrast and T2 modalities to train ML models to differentiate HCC from ICC, reaching an accuracy of 73% ^43^. Oestmann et al. ^44^ constructed a 3D-CNN model using a contrast-enhanced MRI dataset of 118 patients with 150 lesions, to differentiate between HCCs and non-HCCs which reached an overall accuracy of 87.3%. In the previous investigations, studies used DWI to improve diagnostic accuracy without applying further quantitative feature implementation ^11–13,45^.

Utilizing DWI, a non-invasive technique, in conjunction with radiomics, has the potential to significantly contribute to the precise monitoring of pathology outcomes ^13,24,26,46,47^. We conducted this two-center study using machine-learning approaches combined with radiomics features extracted from DWI, minimizing the uncertainty of decisions in LI-RADS categorization.

Selected significant GLCM features, Wavelet-LHH-IDM and Wavelet-HLH-IDMN, provide insight into the local homogeneity of pixel intensities in various orientations within the wavelet-transformed image, offering sensitivity across different intensity scales. Similarly, Wavelet-HLL- Joint Average focuses on specific frequency components in both horizontal and vertical directions, shedding light on the average relationship between pixel intensities at different scales and orientations. Additionally, selected shape-based radiomic features highlighted that irregular shapes or asymmetry in tumors may correlate with more aggressive behavior ^48,49^.

Major Axis, Elongation, and Sphericity metrics contribute valuable information about lesion extent, elongation, and overall geometry, respectively. In the context of selected first-order features, interpretations involve understanding tissue characteristics and their potential associations with pathophysiological changes, considering alterations in cellularity, necrosis, microstructural properties, and vascularity. Furthermore, selected GLSZM features quantify size and intensity variations of homogeneous regions, while selected GLDM features capture changes in cellularity, tissue heterogeneity, and microstructural alterations, providing insights into spatial dependencies, homogeneity, and intricate textural patterns associated with specific tumor characteristics or microstructural organization ^49^.

The results indicated that the designed SVM classifier could reliably and accurately diagnose benign and HCC classes with accuracy/sensitivity/specificity of 0.92/0.94/0.86, respectively. While the RF, MLP, LR, and GB models consistently achieved high accuracy during training on 3T images, as evidenced by ROC curves across folds, a significant divergence emerged when evaluating their performance on the test dataset encompassing both 1.5T and 3T images. SVM’s ability to maintain accuracy on the test dataset positions it as a promising candidate for real-world applications. Potential overfitting, model complexity, or sensitivity to specific characteristics in the training data might contribute to the observed disparities. It is crucial to carefully examine the training metrics and the behavior of the model on separate datasets to guarantee dependable and consistent performance in real-world scenarios.

Our study had several limitations. First, our training and testing sample size was relatively small when compared to the extensive databases typically relied upon for translating results to clinical settings. This could potentially affect the generalizability of our findings. To address this issue, we assessed the technique’s generalizability by utilizing an external dataset obtained from a different MRI unit. Second, selection bias may have existed because we retrospectively analyzed patients with cirrhosis without prior treatment. Pathologic assessment was limited, as not all patients underwent biopsy or liver transplantation. We also excluded some lesions without pathology or follow-up imaging, as malignancy/DWI, or precision-matching between the MRI images and histopathologic findings could not be performed.

## Conclusion

Our study explored machine learning models using radiomics features in classifying HCC from benign nodules by extracting water restriction patterns. By focusing on the distribution of water restriction and selecting important features, we achieved an accurate classification of HCC from benign nodules with 92% accuracy. For future studies, it is essential to establish a correlation between MRI features and histopathological markers employed in primary tumor evaluation to deepen our understanding of cancer progression in HCC and metastatic stages. Importantly, these prediction models may significantly impact clinical decision-making regarding systematic liver biopsy in lesions with uncertain MRI findings in the early stages.

## Abbreviations

HCC: Hepatocellular carcinoma
LI-RADS: Liver Imaging Reporting and Data System
DCE: Dynamic contrast-enhanced
DWI: Diffusion-weighted imaging
ADC: Apparent diffusion coefficient
ICC: Intrahepatic cholangiocarcinoma
EPI: Echo planar imaging
SPGR: Spoiled gradient-recalled echo
SSFSE: Single-shot fast spin echo
ROI: Region of interest
GLCM: Grey-level co-occurrence matrix
GLSZM: Gray level size zone matrix
GLRLM: Gray level run length matrix
GLDM: Gray level dependence matrix
RF: Random Forest
LR: Logistic regression
SVM: Support vector machine
MLP: Multilayer perceptron
GB: Gradient Boosting
ROC: Receiver operating characteristic
AUC: Area-under-curve

## Data Availability

All data produced in the present study are available upon reasonable request to the authors

## References

1. Sung H, Ferlay J, Siegel RL, et al. Global Cancer Statistics 2020: GLOBOCAN Estimates of Incidence and Mortality Worldwide for 36 Cancers in 185 Countries. CA Cancer J Clin. May 2021;71(3):209-249. doi:10.3322/caac.21660

2. Caraiani C-N, Marian D, Militaru C, Calin A, Badea R. The role of the diffusion sequence in magnetic resonance imaging for the differential diagnosis between hepatocellular carcinoma and benign liver lesions. Clujul Medical. 2016;89(2):241.

3. Inchingolo R, De Gaetano AM, Curione D, et al. Role of diffusion-weighted imaging, apparent diffusion coefficient and correlation with hepatobiliary phase findings in the differentiation of hepatocellular carcinoma from dysplastic nodules in cirrhotic liver. European radiology. 2015;25(4):1087–1096.

4. Llovet JM, Kelley RK, Villanueva A, et al. Hepatocellular carcinoma. Nature Reviews Disease Primers. 2021/01/21 2021;7(1):6. doi:10.1038/s41572-020-00240-3

5. Nalaini F, Shahbazi F, Mousavinezhad SM, Ansari A, Salehi M. Diagnostic accuracy of apparent diffusion coefficient (ADC) value in differentiating malignant from benign solid liver lesions: a systematic review and meta-analysis. The British Journal of Radiology. 2021;94:20210059.

6. Zhong X, Tang H, Guan T, et al. Added Value of Quantitative Apparent Diffusion Coefficients for Identifying Small Hepatocellular Carcinoma from Benign Nodule Categorized as LI-RADS 3 and 4 in Cirrhosis. J Clin Transl Hepatol. Feb 28 2022;10(1):34–41. doi:10.14218/jcth.2021.00053

7. Jiang HY, Chen J, Xia CC, Cao LK, Duan T, Song B. Noninvasive imaging of hepatocellular carcinoma: From diagnosis to prognosis. World J Gastroenterol. Jun 14 2018;24(22):2348–2362. doi:10.3748/wjg.v24.i22.2348

8. Wu H, Liang Y, Jiang X, et al. Meta-analysis of intravoxel incoherent motion magnetic resonance imaging in differentiating focal lesions of the liver. Medicine (Baltimore). 2018/08// 2018;97(34):e12071. doi:10.1097/md.0000000000012071

9. Messina C, Bignone R, Bruno A, et al. Diffusion-Weighted Imaging in Oncology: An Update. Cancers (Basel). 2020;12(6). doi:10.3390/cancers12061493 Accessed 2020/06//.

10. Zarghampour M, Fouladi DF, Pandey A, et al. Utility of volumetric contrast-enhanced and diffusion-weighted MRI in differentiating between common primary hypervascular liver tumors. J Magn Reson Imaging. 2018/10// 2018;48(4):1080-1090. doi:10.1002/jmri.26032

11. Peng J, Li J-J, Li J, et al. Could ADC values be a promising diagnostic criterion for differentiating malignant and benign hepatic lesions in Asian populations: A meta-analysis. Medicine (Baltimore). 2016/11// 2016;95(48):e5470. doi:10.1097/md.0000000000005470

12. Yang D, Zhang J, Han D, Jin E, Yang Z. The role of apparent diffusion coefficient values in characterization of solid focal liver lesions: a prospective and comparative clinical study. Sci China Life Sci. 2017/01// 2017;60(1):16-22. doi:10.1007/s11427-016-0387-4

13. Zhong X, Guan T, Tang D, et al. Differentiation of small (≤ 3 cm) hepatocellular carcinomas from benign nodules in cirrhotic liver: the added additive value of MRI-based radiomics analysis to LI-RADS version 2018 algorithm. BMC gastroenterology. 2021;21(1):1–10.

14. Wang YXJ, Huang H, Zheng CJ, Xiao BH, Chevallier O, Wang W. Diffusion-weighted MRI of the liver: challenges and some solutions for the quantification of apparent diffusion coefficient and intravoxel incoherent motion. Am J Nucl Med Mol Imaging. 2021;11(2):107–142.

15. Abugamra S, Yassin A, Abdel-Rehim ASM, Sheha DS. Apparent diffusion coefficient for differentiating between benign and malignant hepatic focal lesions. Egyptian Liver Journal. 2020/12/10 2020;10(1):57. doi:10.1186/s43066-020-00068-2

16. Surov A, Pech M, Omari J, et al. Diffusion-Weighted Imaging Reflects Tumor Grading and Microvascular Invasion in Hepatocellular Carcinoma. Liver Cancer. Feb 2021;10(1):10–24. doi:10.1159/000511384

17. Pankaj Jain T, Kan WT, Edward S, Fernon H, Kansan Naider R. Evaluation of ADCratio on liver MRI diffusion to discriminate benign versus malignant solid liver lesions. European Journal of Radiology Open. 2018/01/01/ 2018;5:209-214. 10.1016/j.ejro.2018.10.002

18. Ronot M, Purcell Y, Vilgrain V. Hepatocellular Carcinoma: Current Imaging Modalities for Diagnosis and Prognosis. Dig Dis Sci. Apr 2019;64(4):934–950. doi:10.1007/s10620-019-05547-0

19. Liu Z, Wang S, Di Dong JW, et al. The applications of radiomics in precision diagnosis and treatment of oncology: opportunities and challenges. Theranostics. 2019;9(5):1303.

20. Liu Z, Wang S, Dong D, et al. The Applications of Radiomics in Precision Diagnosis and Treatment of Oncology: Opportunities and Challenges. Theranostics. 2019;9(5):1303–1322. doi:10.7150/thno.30309

21. Shur JD, Doran SJ, Kumar S, et al. Radiomics in Oncology: A Practical Guide. RadioGraphics. 2021;41(6):1717–1732. doi:10.1148/rg.2021210037

22. Zhang B, Tian J, Dong D, et al. Radiomics Features of Multiparametric MRI as Novel Prognostic Factors in Advanced Nasopharyngeal Carcinoma. Clinical Cancer Research. 2017;23(15):4259–4269. doi:10.1158/1078-0432.Ccr-16-2910

23. Bera K, Braman N, Gupta A, Velcheti V, Madabhushi A. Predicting cancer outcomes with radiomics and artificial intelligence in radiology. Nature Reviews Clinical Oncology. 2022/02/01 2022;19(2):132-146. doi:10.1038/s41571-021-00560-7

24. Lewis S, Peti S, Hectors SJ, et al. Volumetric quantitative histogram analysis using diffusion-weighted magnetic resonance imaging to differentiate HCC from other primary liver cancers. Abdominal Radiology. 2019/03/01 2019;44(3):912-922. doi:10.1007/s00261-019-01906-7

25. Alksas A, Shehata M, Saleh GA, et al. A novel computer-aided diagnostic system for accurate detection and grading of liver tumors. Scientific reports. 2021;11(1):1–18.

26. Mokrane FZ, Lu L, Vavasseur A, et al. Radiomics machine-learning signature for diagnosis of hepatocellular carcinoma in cirrhotic patients with indeterminate liver nodules. Eur Radiol. Jan 2020;30(1):558–570. doi:10.1007/s00330-019-06347-w

27. Xia T, Zhao B, Li B, et al. MRI-Based Radiomics and Deep Learning in Biological Characteristics and Prognosis of Hepatocellular Carcinoma: Opportunities and Challenges. Journal of Magnetic Resonance Imaging. 2024;59(3):767–783.

28. Xia T, Zhao B, Li B, et al. MRI-Based Radiomics and Deep Learning in Biological Characteristics and Prognosis of Hepatocellular Carcinoma: Opportunities and Challenges. Journal of Magnetic Resonance Imaging. 2024;59(3):767–783. 10.1002/jmri.28982

29. Dong X, Yang J, Zhang B, et al. Deep Learning Radiomics Model of Dynamic Contrast-Enhanced MRI for Evaluating Vessels Encapsulating Tumor Clusters and Prognosis in Hepatocellular Carcinoma. Journal of Magnetic Resonance Imaging. 2024;59(1):108–119. 10.1002/jmri.28745

30. Zhou Y, Zhou G, Zhang J, Xu C, Zhu F, Xu P. DCE-MRI based radiomics nomogram for preoperatively differentiating combined hepatocellular-cholangiocarcinoma from mass-forming intrahepatic cholangiocarcinoma. European Radiology. 2022/07/01 2022;32(7):5004-5015. doi:10.1007/s00330-022-08548-2

31. Zheng J, Du P-Z, Yang C, et al. DCE-MRI-based radiomics in predicting angiopoietin-2 expression in hepatocellular carcinoma. Abdominal Radiology. 2023/11/01 2023;48(11):3343-3352. doi:10.1007/s00261-023-04007-8

32. Keenan KE, Delfino JG, Jordanova KV, et al. Challenges in ensuring the generalizability of image quantitation methods for MRI. Medical Physics. 2022;49(4):2820–2835. 10.1002/mp.15195

33. Shukla-Dave A, Obuchowski NA, Chenevert TL, et al. Quantitative imaging biomarkers alliance (QIBA) recommendations for improved precision of DWI and DCE-MRI derived biomarkers in multicenter oncology trials. Journal of Magnetic Resonance Imaging. 2019;49(7):e101–e121. 10.1002/jmri.26518

34. Chernyak V, Fowler KJ, Kamaya A, et al. Liver Imaging Reporting and Data System (LI-RADS) version 2018: imaging of hepatocellular carcinoma in at-risk patients. Radiology. 2018;289(3):816–830.

35. van Griethuysen JJM, Fedorov A, Parmar C, et al. Computational Radiomics System to Decode the Radiographic Phenotype. Cancer Res. Nov 1 2017;77(21):e104–e107. doi:10.1158/0008-5472.Can-17-0339

36. Pedregosa F, Varoquaux G, Gramfort A, et al. Scikit-learn: Machine learning in Python. the Journal of machine Learning research. 2011;12:2825–2830.

37. Joo I, Lee JM, Yoon JH. Imaging diagnosis of intrahepatic and perihilar cholangiocarcinoma: recent advances and challenges. Radiology. 2018;288(1):7–13.

38. Kim JH, Joo I, Lee JM. Atypical appearance of hepatocellular carcinoma and its mimickers: how to solve challenging cases using gadoxetic acid-enhanced liver magnetic resonance imaging. Korean Journal of Radiology. 2019;20(7):1019–1041.

39. Cerny M, Chernyak V, Olivié D, et al. LI-RADS version 2018 ancillary features at MRI. Radiographics. 2018;38(7):1973-2001.

40. Fotouhi M, Samadi Khoshe Mehr F, Delazar S, et al. Assessment of LI-RADS efficacy in classification of hepatocellular carcinoma and benign liver nodules using DCE-MRI features and machine learning. European Journal of Radiology Open. 2023/12/01/ 2023;11:100535. 10.1016/j.ejro.2023.100535

41. Arian A, Abdullah AD, Taher HJ, Suhail Alareer H, Fotouhi M. Diagnostic Values of the Liver Imaging Reporting and Data System in the Detection and Characterization of Hepatocellular Carcinoma: A Systematic Review and Meta-Analysis. Cureus. Mar 2023;15(3):e36082. doi:10.7759/cureus.36082

42. Liu HF, Lu Y, Wang Q, Lu YJ, Xing W. Machine Learning-Based CEMRI Radiomics Integrating LI-RADS Features Achieves Optimal Evaluation of Hepatocellular Carcinoma Differentiation. J Hepatocell Carcinoma. 2023;10:2103–2115. doi:10.2147/jhc.S434895

43. Hu R, Li H, Horng H, et al. Automated machine learning for differentiation of hepatocellular carcinoma from intrahepatic cholangiocarcinoma on multiphasic MRI. Scientific Reports. 2022/05/13 2022;12(1):7924. doi:10.1038/s41598-022-11997-w

44. Oestmann PM, Wang CJ, Savic LJ, et al. Deep learning–assisted differentiation of pathologically proven atypical and typical hepatocellular carcinoma (HCC) versus non-HCC on contrast-enhanced MRI of the liver. European Radiology. 2021/07/01 2021;31(7):4981-4990. doi:10.1007/s00330-020-07559-1

45. Yazdi NA, Dashti H, Safaei M, et al. Radiologic and Pathologic Findings of a Huge Solitary Fibrous Tumor of the Liver with Malignant Transformation: A Case Report. Iranian Journal of Radiology. 2020;17(1)

46. Jiang H, Liu X, Chen J, et al. Man or machine? Prospective comparison of the version 2018 EASL, LI-RADS criteria and a radiomics model to diagnose hepatocellular carcinoma. Cancer Imaging. 2019/12/05 2019;19(1):84. doi:10.1186/s40644-019-0266-9

47. Bera K, Braman N, Gupta A, Velcheti V, Madabhushi A. Predicting cancer outcomes with radiomics and artificial intelligence in radiology. Nat Rev Clin Oncol. Feb 2022;19(2):132–146. doi:10.1038/s41571-021-00560-7

48. Miranda J, Horvat N, Fonseca GM, et al. Current status and future perspectives of radiomics in hepatocellular carcinoma. World J Gastroenterol. Jan 7 2023;29(1):43–60. doi:10.3748/wjg.v29.i1.43

49. Shur JD, Doran SJ, Kumar S, et al. Radiomics in Oncology: A Practical Guide. RadioGraphics. 2021/10/01 2021;41(6):1717-1732. doi:10.1148/rg.2021210037

